# A framework for identifying regional outbreak and spread of COVID-19 from one-minute population-wide surveys

**DOI:** 10.1101/2020.03.19.20038844

**Authors:** Hagai Rossman, Ayya Keshet, Smadar Shilo, Amir Gavrieli, Tal Bauman, Ori Cohen, Ran Balicer, Benjamin Geiger, Yuval Dor, Eran Segal

## Abstract

Coronavirus infection spreads in clusters and therefore early identification of these clusters is critical for slowing down the spread of the virus. Here, we propose that daily population-wide surveys that assess the development of symptoms caused by the virus could serve as a strategic and valuable tool for identifying such clusters to inform epidemiologists, public health officials, and policy makers. We show preliminary results from a survey of over 58,000 Israelis and call for an international consortium to extend this concept in order to develop predictive models. We expect such data to allow: Faster detection of spreading zones and patients; Obtaining a current snapshot of the number of people in each area who have developed symptoms; Predicting future spreading zones several days before an outbreak occurs; Evaluating the effectiveness of the various social distancing measures taken, and their contribution to reduce the number of symptomatic people. Such information can provide a valuable tool for decision makers to decide which areas need strengthening of social distancing measures and which areas can be relieved. Preliminary analysis shows that in neighborhoods with confirmed COVID-19 patient history, more responders report on COVID-19 associated symptoms, demonstrating the potential utility of our approach for detection of outbreaks. Researchers from other countries including the U.S, India, Italy, Spain, Germany, Mexico, Finland, Sweden, Norway and several others have adopted our approach and we are collaborating to further improve it. We call with urgency for other countries to join this international consortium, and to share methods and data collected from these daily, simple, one-minute surveys.

---

In December 2019 a novel coronavirus was isolated, after a cluster of patients with pneumonia of unknown cause were diagnosed in China. This new isolate was named 2019-nCoV and is the cause for the COVID-19 disease (Na Zhu et al. 2019). The virus has led to an ongoing outbreak and an unprecedented international health crisis. Update reports of the World Health Organization (WHO) state that at the moment there are at the low hundreds of thousands confirmed cases and thousands of deaths of patients who were infected by COVID-19 globally ^1^. This number is rapidly increasing and most probably is a vast underestimation of the real number of patients worldwide, as infected individuals are contagious even when minimally symptomatic or asymptomatic ^2^. The spread of the disease has presented an extreme challenge to the international community. Although the WHO issues health policies recommendations, many policy-makers from different countries have chosen and implemented different strategies. These strategies depend on many factors, including the local spread of COVID-19, healthcare system resources, economical and political factors, public adherence and their perception of the situation.

In Israel, the first infection of COVID-19 was confirmed on February 21st 2020, and in response, the Israeli Ministry of Health (MOH) instructed individuals who returned to Israel from specific countries, in which COVID-19 was spreading, to go into a 14-day home isolation. Since then, Israel has gradually imposed several measures aimed at slowing the spread of the coronavirus (Figure 2). On March 9th, 14-day home isolation was extended to individuals arriving from anywhere in the world starting from their date of arrival. Individuals that were in close contact, defined by being within approximately 2 meters (6 ft 7 in) from a COVID-19 patient for more than 15 minutes, were also instructed to be in home isolation for a similar time period. Symptomatic individuals with fever above 38 Celsius and respiratory symptoms (cough or shortness of breath) were instructed to stay home for two days after the fever has dropped down ^3^. On March 11th, gatherings were limited to a maximum of 100 people and on March 15th, to 10 people, with attendees advised to keep a minimal distance of 2 m (6 ft 7 in) between one another. On March 19th, a national state of emergency was declared in the country, and Israeli citizens were banned from leaving their homes unless absolutely necessary with the exception of essential services that remained open. On March 20st, the first death of an Isreali citizen as a result of COVID-19 infection occurred.

**Figure 1:**
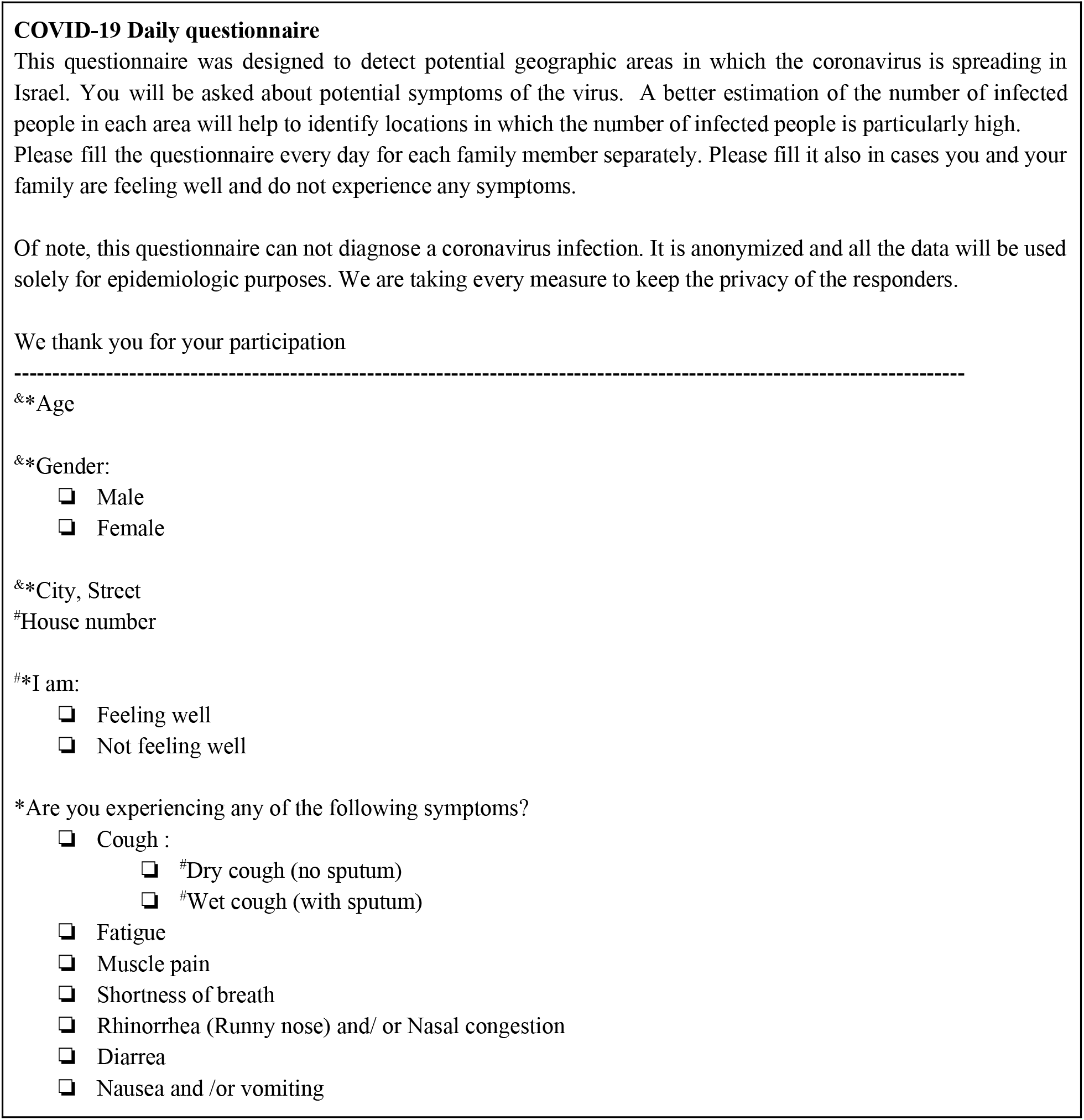
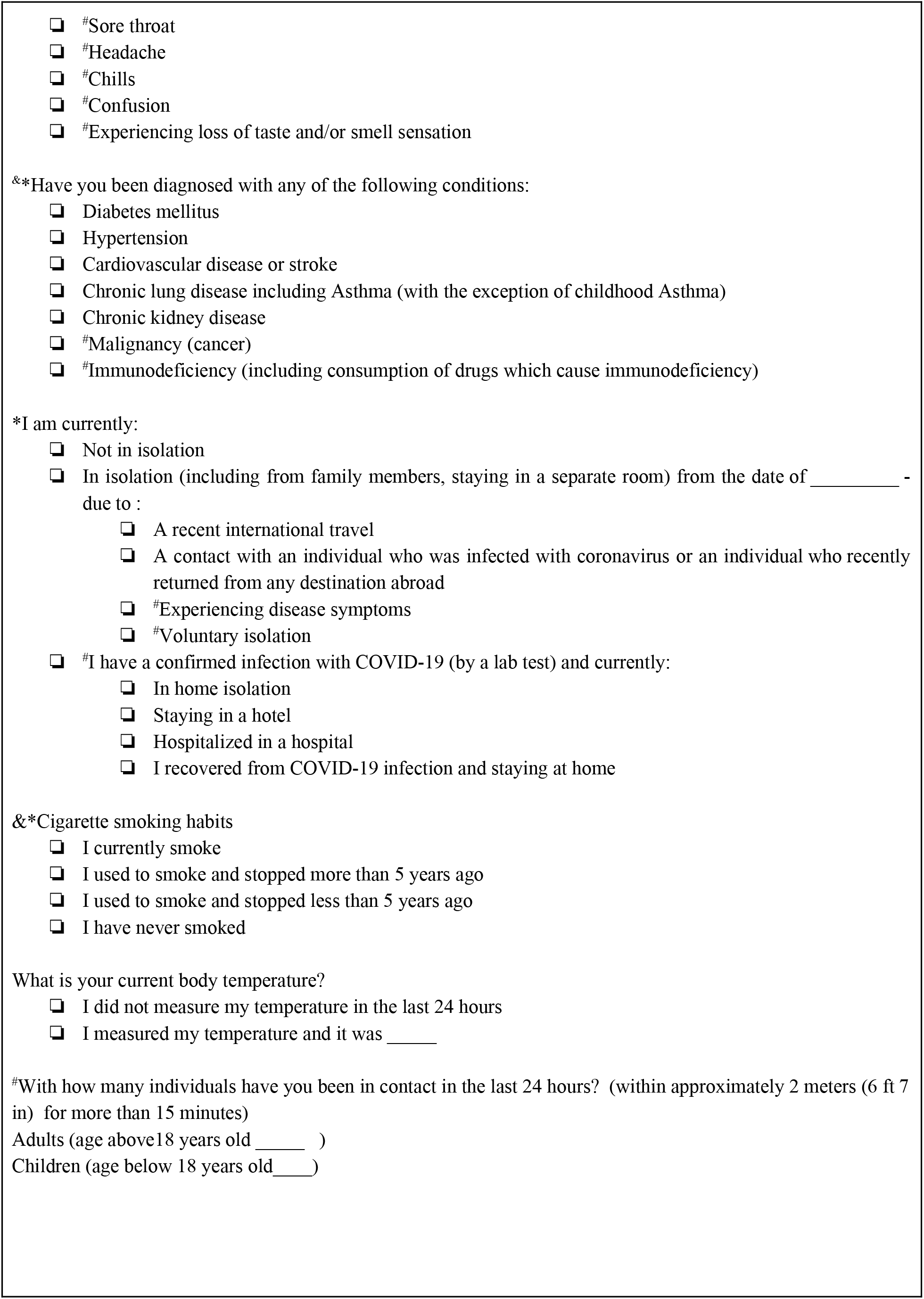
COVID-19 Daily questionnaire *Questions that the responder is required to answer, ^&^ Questions that should be filled only once, ^#^Questions that were added in the new version of the questionnaire and are therefore not analysed in this paper

**Figure 2:**
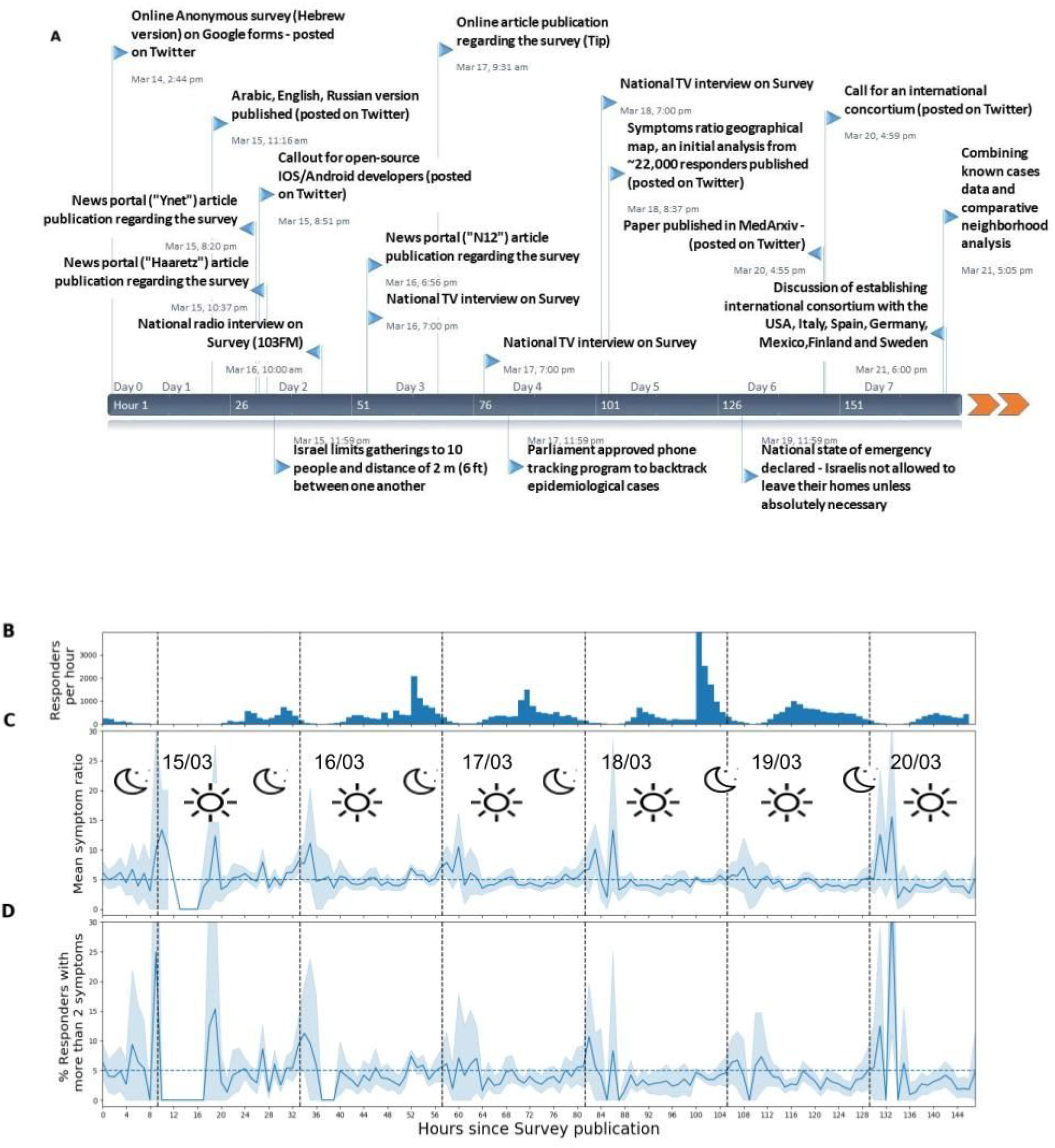
**A:** Project timeline describing all major events in its development including national events which affected and drove its process, from time survey online publication (March 14th, 14:44) to March 21, 18:00 **B**: Number of responding participants per hour. **C**: Mean symptoms ratio of responders. **D**: Percentage of responders who reported more than two symptoms.

One of the major challenges of the current pandemic to date is disease detection and diagnosis. While the gold standard for COVID-19 diagnosis is detecting the virus by a real-time RT-PCR testing ^4^, current resource and policy limitations in many countries restrict the amount of testing that can be performed. On March 12, 600 tests by 4 different laboratories were being done across Israel ^5^. Although the number of tests per day is gradually growing, these cannot provide a full snapshot of the spread of the virus, especially since the MOHs guidelines are to test only individuals who were in close contact with a confirmed case.

As testing the entire population for the presence of 2019-nCoVis is currently not feasible, we developed a simple one-minute online questionnaire with the goal of early and temporal detection of geographic clusters in which the virus is spreading. The survey was posted online as a Google Form (http://predict-corona.org/) on March 14th. This additional information provides real-time analysis of symptoms over the course of days as the virus spreads through the population, and renders an informative situation snapshot.

The survey contains questions on age, gender, geographic location (city, street, zip code), isolation status and smoking habits. Furthermore, responders were asked to report whether they experience symptoms which were defined as common symptoms of COVID-19 by healthcare professionals based on the existing literature ^6^. Several other symptoms which are not common in patients with COVID-19 infection but are common in other infectious diseases were also included to discern possible COVID-19 patients.. The symptoms that were included are cough, fatigue, myalgia (muscle pain), shortness of breath, rhinorrhea or nasal congestion, diarrhea and nausea or vomiting. In addition, participants were asked about the existence of one of the following chronic health conditions: Diabetes mellitus, Hypertension, Ischemic heart disease, Asthma, chronic lung disease and chronic kidney disease. Participants were also asked to measure their daily body temperature and document it in the questionnaire (Figure 1).

For each responder we calculate the *symptoms ratio*:

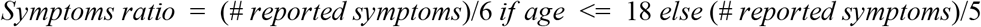

The number of reported symptoms divided by the number of symptoms in our predefined list. Symptoms in this list were predefined by the Israeli MOH and included: shortness of breath, fatigue, cough, muscle pains and a high fever (body temperature of over 38 degrees celsius). For responders under the age of 18 - nausea or vomiting was also included in the calculation.

Responders were associated with an area in Israel using the address provided to create a colormap of Israel by the aggregated symptoms ratio as defined by the MOH.

We made an effort to distribute and reach all sectors of Israel’s population. The survey is distributed in 5 languages - Hebrew, Arabic, English, Russian, and Amharic to reflect and serve the diverse spoken languages of Israel’s population. Although at this point we do not see an unbiased representation of the Israeli population in our responses, special efforts to reach underrepresented populations are currently done through several channels: call centers, media appearance and promotion of the survey through Arabic-speaking TV stations to gain interest and compliance in all sectors of the population.

The questionnaire was first distributed online on March 14th 2020 at 14:43 (Israeli time zone, GMT+2) and was spread via social media and press. To this date, March 21st, 18pm, there have been 54,059 responses, including 50,572 (93.55%) adults and 3,488 (6.45%) children. The characteristics of the responders are described in Table 1, and by response date in Table S1 in the Appendix. Altogether, 2,216 (4.1%) responders reported that they are currently in isolation. For those reported as being in isolation, 1,116 (50.36%) responders were due to a recent international travel and 1,100 (49.63%) were due to a contact with an individual who was infected with COVID-19 or an individual who recently returned from any destination abroad. A new version of the questionnaire was established on March 21 driven by new policies that have been implemented by the ministry of health (Figure 2) and a comprehensive meta-analysis of 14 published papers on patients with COVID-19 symptoms (7 of which were on patients over 18 years old and 7 on patients under). By that time, new data was published which we relied on instead ^6^. This version includes several more questions (Figure 1) and was not distributed yet.

**Table 1.** Baseline characteristics of questionnaire responders.

| **Table 1. Baseline characteristics of questionnaire responders** |  |  |  |  |  |
| --- | --- | --- | --- | --- | --- |
| **Characteristic, mean (SD) or %** | **All**<br>**(n = 54,059)** | **Not in home isolation**<br>**(n = 51,843)**<br>**(95.9%)** | **In home isolation**<br>**(n = 2,216)**<br>**(4.1%)** | **Adults**<br>**(n = 50,572)**<br>**(93.55%)** | **Children**<br>**- under 18**<br>**(n = 3,488)**<br>**(6.45%)** |
| Age (years) | 44.93 (18.13) | 45.07 (18.14) | 41.59 (17.56) | 47.21 (16.4) | 11.88 (5.22) |
| Sex - Male | 25,196 (46.61%) | 24,100 (46.49%) | 1,096 (49.46%) | 23,446 (46.36%) | 1,750 (50.17%) |
| Smoking history<br>(previously smoked/<br>currently smoking) | 19,484 (36.04%) | 18,742 (36.15%) | 742 (33.48%) | 19,343 (38.25%) | 141 (4.04%) |
| Presence of a chronic<br>medical conditions | 10,646 (19.69%) | 10,308 (19.88%) | 338 (15.25%) | 10,459 (20.68%) | 187 (5.36%) |
| **Symptoms** |  |  |  |  |  |
| No symptoms (Feel<br>good) | 44,850 (82.96%) | 43,086 (83.11%) | 1,764 (79.6%) | 42,166 (83.38%) | 2,684 (76.95%) |
| Body temperature<br>(Celcius) | 36.5 (0.45) | 36.5 (0.45) | 36.54 (0.47) | 36.49 (0.43) | 36.64 (0.67) |
| Body temperature<br>equals or above 38<br>degrees celsius | 173 (0.32%) | 151 (0.29%) | 22 (0.99%) | 94 (0.19%) | 79 (2.26%) |
| Nausea and vomiting | 326 (0.6%) | 306 (0.59%) | 20 (0.9%) | 282 (0.56%) | 44 (1.26%) |
| Myalgia (Muscle pain) | 1,737 (3.21%) | 1,637 (3.16%) | 100 (4.51%) | 1,635 (3.23%) | 102 (2.92%) |
| Rhinorrhea or nasal<br>congestion | 7,336 (13.57%) | 7,027 (13.55%) | 309 (13.94%) | 6,585 (13.02%) | 751 (21.53%) |
| Fatigue | 2,272 (4.2%) | 2,142 (4.13%) | 130 (5.87%) | 2,113 (4.18%) | 159 (4.56%) |
| Shortness of breath | 1,042 (1.93%) | 973 (1.88%) | 69 (3.11%) | 980 (1.94%) | 62 (1.78%) |
| Cough | 7,552 (13.97%) | 7,183 (13.86%) | 369 (16.65%) | 6,906 (13.66%) | 646 (18.52%) |
| Diarrhea | 938 (1.74%) | 879 (1.7%) | 59 (2.66%) | 827 (1.64%) | 111 (3.18%) |
| **Symptoms ratio** |  |  |  |  |  |
| Symptoms ratio | 0.05 (0.11) | 0.05 (0.11) | 0.06 (0.14) | 0.05 (0.11) | 0.05 (0.12) |

\* Symptoms ratio was calculated as the number of reported symptoms divided by the number of symptoms in the predefined MOH symptoms list.

We performed several analyses on this preliminary data. First, we analysed the percentage of symptoms by time (Figure 2 Middle). Mean symptoms ratio of responders was roughly stable at 0.05 during daytime, in which the rate of responses was higher. The percentage of responders who reported more than 2 symptoms was also similar in these timeframes. We next analysed the symptoms ratio of responders by geographical locations in Israel (Figure 3). This analysis revealed a different rate of symptoms in responders from different cities and different neighborhoods which are geographically close to each other, which might hint at the ability to detect changes in a high geographical resolution.

**Figure 3:**
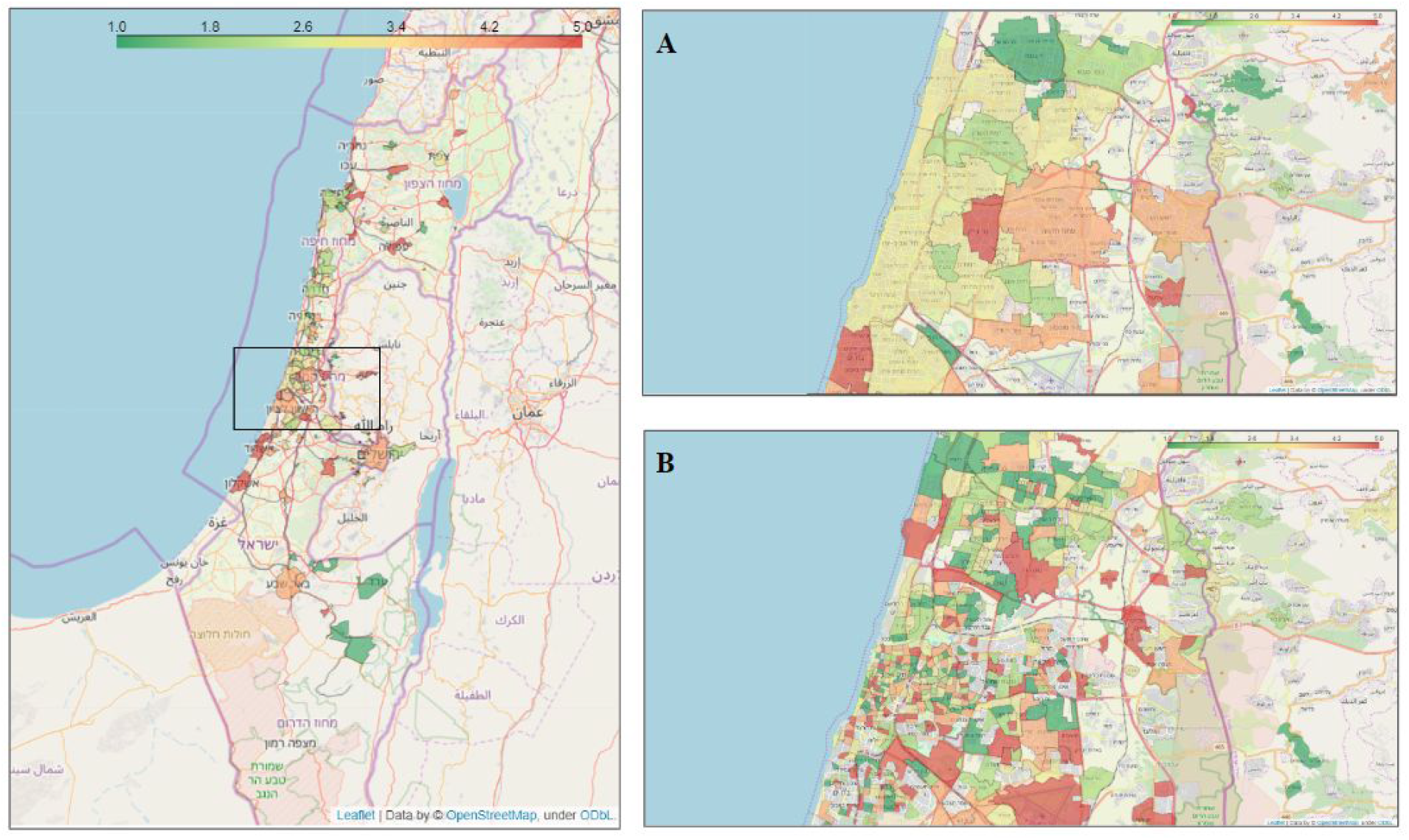
Average COVID-19 associated symptoms region map. City municipal regions with at least 30 responders and neighborhoods with at least 10 responders are shown. Each region is colored by a category defined by the average symptoms ratio, calculated by averaging the reported symptoms rate by responders in that city or neighborhood. Green - low symptoms rate, red - high symptoms rate. **A:** Area of Tel-Aviv and Gush-Dan with city regions. **B:** Area of Tel-Aviv and Gush-Dan with neighborhood regions.

Finally, we analysed the association between symptoms that were reported by the responders, to symptoms described in the literature as characterizing patients with COVID-19. We divided the responders into two groups depending on whether they were living in neighborhoods in which confirmed cases of COVID-19 infection were present or not ^6^ (Figure 4).

**Figure 4:**
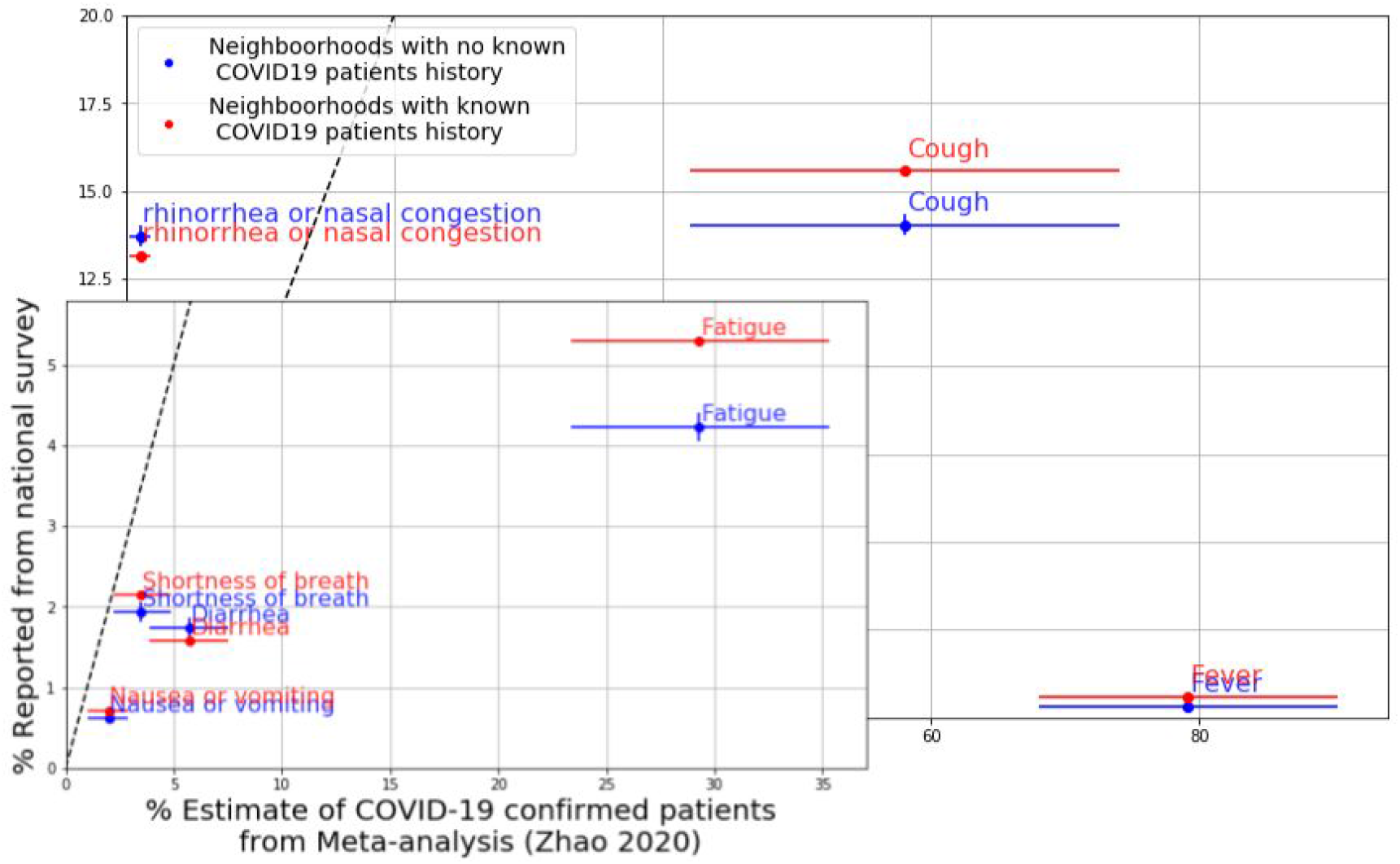
Symptoms prevalence for responders from neighborhoods where confirmed cases were present (red), and for responders from neighborhoods where no confirmed cases were present (blue). X axis - Estimates and 95% confidence intervals of COVID-19 patients from a meta-analysis ^6^, Y axis - Prevalence from survey data, cand bootstrap estimates of 95% confidence intervals. Dashed diagonal line is y=x.

In conclusion, here we developed a short survey based on symptoms associated with COVID-19 infection with the primary goal of early detection of clusters of COVID-19 outbreak. At the time of writing, only seven days after the survey was published, 54,059 responders have already completed it. As expected, we are also detecting a higher percentage of symptoms among individuals who are in home isolation, compared to those who are not (0.06 and 0.05 respectively, *p* < 5*e* − 10). Temporal analysis of the symptoms rate during the first week shows a relatively stable rate in the population at daytime, where response rate is relatively high (Figure 1). Although the spread of COVID-19 infection is exponential ^7^, and the number of confirmed COVID-19 patients in Israel has increased from 193 cases on March 14th 2020 to 883 patients on March 21th noontime ^8^, it has yet to reach the vast majority of Israel’s population. We therefore hypothesize that these symptoms may reflect other respiratory infections which are prevalent in Israel during this period (such as infections caused by an influenza virus), as many of these diseases share common symptoms ^9^. This is supported by the fact that we detected symptoms which were less prevalent in patients with COVID-19 at a similar rate as those who were common in these patients. For example, cough, which is one of the most common symptoms in patients with COVID-19 infection ^6^ was present in a similar percentage of responders as rhinorrhea or nasal congestion which is uncommon in this infection (14%) (Table 1, Figure 4). If our hypothesis is correct, this rate can be viewed as a baseline rate that will allow us to further detect subtle changes with a high geographic resolution which may reflect a spread of COVID-19 infection.

These findings encourage us to reach as much of the population as possible in a minimal amount of time to actively create a reliable baseline of the symptoms for the population. Interestingly, at the moment we detect a higher prevalence of symptoms that are more common in patients with COVID-19 infection such as cough in responders from neighborhoods with known COVID-19 patients compared to symptoms less associated with the disease such as rhinorrhea or nasal congestion which were more present in the other responders (Figure 4).

Our tool has several potential applications. First, while it does not have the ability to diagnose individual cases of COVID-19 infections, it may help predict future spreading zones a few days before an outbreak occurs, with a high level of accuracy given a sufficient sample size. We currently provide a colormap of Israel by regions of symptoms ratio (Figure 3), however, as the daily response rate will increase, enough data will be collected, enabling us to implement prediction models. These we hope would be leveraged by policy-makers to make informed decisions, by utilization of efficient regional prevention strategies rather than a universal approach. Second, it may also be used to evaluate the effectiveness of prevention strategies implemented by public health organizations, such as the various social distancing measures which are currently being employed in many countries, including Israel. This can be done by measuring the effect of different strategies on reducing the number of symptomatic individuals. Third, it may help in understanding the clinical course of COVID-19 infection by tracking the dynamics of symptoms in the population over time.

Addressing the ongoing needs of the medical and scientific community as well as feedback of policy-makers will drive the directions and the focus of our future work. To improve ease of use by responders and streamline the data collection process, we are also building a designated mobile application which is being finalized as these lines are written. We also hope to resolve privacy issues around location sharing in the future application, which will be used ONLY in an aggregated level and can significantly improve our models, and provide valuable insights on population interactions, adherence and disease spread dynamics.

Although our approach has many possible clinical implications, we have so far encountered several challenges. Asking participants to fill the survey may raise issues of information privacy related to the collection of medical data, to avoid any privacy issues that may occur, our survey is filled anonymously. Moreover, since our survey is anonymous we cannot link the same responder’s daily questionnaires, which can provide an individual trend as we proceed. However, our goal is to provide a macro-management tool which is still feasible. Finally, a main challenge when relying on this data is that it is prone to selection bias. We notice that regions with relatively high response rate are regions associated with higher socioeconomic status. It may also be assumed that people who answer these surveys tend to be more adherent to healthcare protocols and thus less likely to be infected. While some bias may decrease as these surveys become more popular and known and thus better reflect the true population, we intend to model and adjust for different factors such as age and location.

In summary, we present a new tool that has the potential to early detect clusters of COVID-19 infection. This paper and the presented analyses were written in great haste and urgency. We were inspired by the WHO executive director Dr. Michael Ryan words: “*Perfection is the enemy of the good when it comes to emergency management*". We urge other countries to adopt this tool and encourage their population to use these daily, simple, one-minute surveys. We call for an international collaboration, which will allow the sharing of methods and collected data. We also call for the “Tech Giants” Google, Facebook and Twitter to collaborate in this international effort by sharing REGIONAL (not personal) information to help us improve our models.

## Data Availability

Surveys are available at http://predict-corona.org/

## Acknowledgments

We thank Dr. Uri Feinstein for assisting us in defining the surveys, symptoms, and medical conditions. We thank Feng Zhang, Ophir Shalem, Will Allen, Ben Silbermann, Ryan Probasco and David Cheng for insightful discussions and look forward to jointly creating an international consortium with them. We thank Ofer Bartal and the people at Israel Corona Map, Hasadna, Tamar Ben-Ami, Micha Hashkes, Hagay Ben-Shushan, Snir Avraham, Bar Kirel, Ariel Terkeltaub, Dorit Hizi, Adam Kariv, Mushon Zer-Aviv, Noam Kastel, Roy Folkman and Iris Kalka for their contribution to our effort.

## Data availability statement

Survey templates are available at http://predict-corona.org/

## Code availability statement

Analysis code will be released at https://github.com/hrossman/Covid19-Survey

## Supplementary material

**Table S1.** Baseline characteristics of questionnaire responders - by days

|  |  |  |  |  |  |  |  |  |  |
| --- | --- | --- | --- | --- | --- | --- | --- | --- | --- |
| **Characteristic, mean (SD) or %** | **All (n =54,059)** | **2020-03-14 (n = 879 ) (1.63%)** | **2020-03-15 (n = 4,38 ) (8.12%)** | **2020-03-16 (n = 9,658) (17.87%)** | **2020-03-17 (n = 8,928) (16.52%)** | **2020-03-18 (n = 13,713 ) (25.37%)** | **2020-03-19 (n =9,935) (18.38%)** | **2020-03-20 (n = 4,746 ) (8.78%)** | **2020-03-21 (n =1,812) (3.35%)** |
| Age (years) | 44.93 (18.13) | 39.55 (17.76) | 40.21 (15.45) | 42.06 (16.45) | 47.34 (18.63) | 44.59 (18.2) | 46.6 (18.87) | 47.63 (18.87) | 48.67 (18.38) |
| Sex - Male | 25,196 (46.61%) | 399 (45.39%) | 2,411 (54.95%) | 4,576 (47.38%) | 3,999 (44.79%) | 6,303 (45.96%) | 4,419 (44.48%) | 2,194 (46.23%) | 895 (49.39%) |
| Smoking history (previously smoked/ currently smoking) | 19,484 (36.04%) | 303 (34.47%) | 1,473 (33.57%) | 3,494 (36.18%) | 3,259 (36.5%) | 5,095 (37.15%) | 3,580 (36.03%) | 1,594 (33.59%) | 686 (37.86%) |
| Presence of a chronic medical conditions | 10,646 (19.69%) | 139 (15.81%) | 652 (14.86%) | 1,582 (16.38%) | 1,930 (21.62%) | 2,715 (19.8%) | 2,141 (21.55%) | 1,074 (22.63%) | 413 (22.79%) |
| No symptoms (Feel good) | 44,850 (82.96%) | 692 (78.73%) | 3,493 (79.6%) | 7,840 (81.18%) | 7,477 (83.75%) | 11,350 (82.77%) | 8,331 (83.86%) | 4,075 (85.86%) | 1,592 (87.86%) |
| Body temperature (Celcius) | 36.5 (0.45) | 36.56 (0.44) | 36.55 (0.43) | 36.55 (0.45) | 36.52 (0.44) | 36.49 (0.45) | 36.47 (0.45) | 36.45 (0.46) | 36.43 (0.43) |
| Body temperature equals or above 38 degrees celsius | 173 (0.32%) | 2 (0.23%) | 17 (0.39%) | 44 (0.46%) | 25 (0.28%) | 44 (0.32%) | 28 (0.28%) | 13 (0.27%) | 0 (0.0%) |
| Nausea and vomiting | 326 (0.6%) | 8 (0.91%) | 32 (0.73%) | 65 (0.67%) | 50 (0.56%) | 75 (0.55%) | 65 (0.65%) | 22 (0.46%) | 9 (0.5%) |
| Myalgia (Muscle pain) | 1,737 (3.21%) | 29 (3.3%) | 165 (3.76%) | 371 (3.84%) | 271 (3.04%) | 448 (3.27%) | 283 (2.85%) | 137 (2.89%) | 33 (1.82%) |
| Rhinorrhea or nasal congestion | 7,336 (13.57%) | 155 (17.63%) | 701 (15.98%) | 1,391 (14.4%) | 1,190 (13.33%) | 1,773 (12.93%) | 1,322 (13.31%) | 582 (12.26%) | 222 (12.25%) |
| Fatigue | 2,272 (4.2%) | 56 (6.37%) | 222 (5.06%) | 494 (5.11%) | 362 (4.05%) | 539 (3.93%) | 378 (3.8%) | 166 (3.5%) | 55 (3.04%) |
| Shortness of breath | 1,042 (1.93%) | 18 (2.05%) | 99 (2.26%) | 231 (2.39%) | 177 (1.98%) | 257 (1.87%) | 149 (1.5%) | 84 (1.77%) | 27 (1.49%) |
| Cough | 7,552 (13.97%) | 142 (16.15%) | 661 (15.06%) | 1,456 (15.08%) | 1,189 (13.32%) | 1,999 (14.58%) | 1,298 (13.06%) | 584 (12.31%) | 223 (12.31%) |
| Diarrhea | 938 (1.74%) | 24 (2.73%) | 95 (2.16%) | 190 (1.97%) | 152 (1.7%) | 229 (1.67%) | 146 (1.47%) | 82 (1.73%) | 20 (1.1%) |
| **Symptoms ratio** |  |  |  |  |  |  |  |  |  |
| Symptoms ratio* | 0.05 (0.11) | 0.06 (0.13) | 0.05 (0.12) | 0.05 (0.12) | 0.05 (0.11) | 0.05 (0.11) | 0.04 (0.11) | 0.04 (0.11) | 0.04 (0.1) |

\* Symptoms ratio was calculated as the number of reported symptoms divided by the number of symptoms in the predefined MOH symptoms list.

